# Aspirin in patients admitted to hospital with COVID-19 (RECOVERY): a randomised, controlled, open-label, platform trial

**DOI:** 10.1101/2021.06.08.21258132

**Authors:** RECOVERY Collaborative Group, Peter W Horby, Guilherme Pessoa-Amorim, Natalie Staplin, Jonathan R Emberson, Mark Campbell, Enti Spata, Leon Peto, Nigel J Brunskill, Simon Tiberi, Victor Chew, Thomas Brown, Hasan Tahir, Beate Ebert, David Chadwick, Tony Whitehouse, Rahuldeb Sarkar, Clive Graham, J Kenneth Baillie, Buddha Basnyat, Maya H Buch, Lucy C Chappell, Jeremy Day, Saul N Faust, Raph L Hamers, Thomas Jaki, Edmund Juszczak, Katie Jeffery, Wei Shen Lim, Alan Montgomery, Andrew Mumford, Kathryn Rowan, Guy Thwaites, Marion Mafham, Richard Haynes, Martin J Landray

## Abstract

**Background:** Aspirin has been proposed as a treatment for COVID-19 on the basis of its antithrombotic properties.

**Methods:** In this randomised, controlled, open-label platform trial, several possible treatments were compared with usual care in patients hospitalised with COVID-19. Eligible and consenting adults were randomly allocated in a 1:1 ratio to either usual standard of care plus 150mg aspirin once daily until discharge or usual standard of care alone using web-based simple (unstratified) randomisation with allocation concealment. The primary outcome was 28-day mortality. The trial is registered with ISRCTN (50189673) and clinicaltrials.gov (NCT04381936).

**Findings:** Between 01 November 2020 and 21 March 2021, 7351 patients were randomly allocated to receive aspirin and 7541 patients to receive usual care alone. Overall, 1222 (17%) patients allocated to aspirin and 1299 (17%) patients allocated to usual care died within 28 days (rate ratio 0·96; 95% confidence interval [CI] 0·89-1·04; p=0·35). Consistent results were seen in all pre-specified subgroups of patients. Patients allocated to aspirin had a slightly shorter duration of hospitalisation (median 8 vs. 9 days) and a higher proportion were discharged from hospital alive within 28 days (75% vs. 74%; rate ratio 1·06; 95% CI 1·02-1·10; p=0·0062). Among those not on invasive mechanical ventilation at baseline, there was no significant difference in the proportion meeting the composite endpoint of invasive mechanical ventilation or death (21% vs. 22%; risk ratio 0·96; 95% CI 0·90-1·03; p=0·23). Aspirin use was associated with an absolute reduction in thrombotic events of 0.6% (SE 0.4%) and an absolute increase in major bleeding events of 0.6% (SE 0.2%).

**Interpretation:** In patients hospitalised with COVID-19, aspirin was not associated with reductions in 28-day mortality or in the risk of progressing to invasive mechanical ventilation or death but was associated with a small increase in the rate of being discharged alive within 28 days.

**Funding:** UK Research and Innovation (Medical Research Council), National Institute of Health Research (Grant ref: MC_PC_19056), and the Wellcome Trust (Grant Ref: 222406/Z/20/Z) through the COVID-19 Therapeutics Accelerator.

## INTRODUCTION

Thrombosis is a key feature of severe COVID-19, with 5-30% of hospitalised patients (depending on illness severity) experiencing a major venous thromboembolic event (mostly pulmonary embolism) and up to 3% an arterial thromboembolic event, particularly myocardial infarction and ischaemic stroke.^1,2^ The risk of thromboembolic complications is reported to be higher in COVID-19 than in other acute medical illnesses and viral respiratory infections, and is associated with worse prognosis.^3,4^

Antiplatelet therapy may have beneficial effects in severe COVID-19 through several mechanisms including inhibition of platelet aggregation, reduction of platelet-derived inflammation, and blocking thrombogenic neutrophil extracellular traps and disseminated intravascular coagulation.^5^ Aspirin is an affordable, globally available drug which at low doses irreversibly inhibits the COX-1 enzyme which is responsible for production of thromboxane A2 and pro-inflammatory prostaglandins. Aspirin can reduce both arterial and venous thrombotic events and has been shown to abolish in-vitro hyperactivity in platelets from SARS-CoV-2 infected patients.^6,7^ Existing randomized evidence has shown that 75-150mg aspirin daily is as effective as higher doses in preventing cardiovascular events.^6^

Seven clinical trials of aspirin in COVID-19 are registered but none have yet reported on the effect of aspirin therapy in COVID-19. Here we report the results of a large randomised controlled trial of aspirin in patients hospitalised with COVID-19.

## METHODS

### Study design and participants

The Randomised Evaluation of COVID-19 therapy (RECOVERY) trial is an investigator-initiated, individually randomised, controlled, open-label, platform trial to evaluate the effects of potential treatments in patients hospitalised with COVID-19. Details of the trial design and results for other treatments evaluated (lopinavir-ritonavir, hydroxychloroquine, dexamethasone, azithromycin, tocilizumab, convalescent plasma, and colchicine) have been published previously.^8-14^ The trial is underway at 177 hospitals in the United Kingdom, two hospitals in Indonesia, and two hospitals in Nepal (appendix pp 5-25), supported in the UK by the National Institute for Health Research Clinical Research Network. The trial is coordinated by the Nuffield Department of Population Health at the University of Oxford (Oxford, UK), the trial sponsor. The trial is conducted in accordance with the principles of the International Conference on Harmonisation–Good Clinical Practice guidelines and approved by the UK Medicines and Healthcare products Regulatory Agency (MHRA) and the Cambridge East Research Ethics Committee (ref: 20/EE/0101). The protocol, statistical analysis plan, and additional information are available on the study website www.recoverytrial.net.

Patients admitted to hospital were eligible for the trial if they had clinically suspected or laboratory confirmed SARS-CoV-2 infection and no medical history that might, in the opinion of the attending clinician, put the patient at significant risk if they were to participate in the trial. Children aged <18 years were not eligible for randomisation to aspirin. Patients with known hypersensitivity to aspirin, a recent history of major bleeding, or currently receiving aspirin or another antiplatelet treatment were excluded. Written informed consent was obtained from all patients, or a legal representative if they were too unwell or unable to provide consent.

### Randomisation and masking

Baseline data were collected using a web-based case report form that included demographics, level of respiratory support, major comorbidities, suitability of the study treatment for a particular patient, and treatment availability at the study site (appendix p33-34). Eligible and consenting adult patients were assigned in a 1:1 ratio to either usual standard of care or usual standard of care plus aspirin using web-based simple (unstratified) randomisation with allocation concealed until after randomisation (appendix p30). For some patients, aspirin was unavailable at the hospital at the time of enrolment or was considered by the managing physician to be either definitely indicated or definitely contraindicated. These patients were excluded from the randomised comparison between usual care plus aspirin and usual care alone. Patients allocated to aspirin were to receive 150 mg by mouth (or nasogastric tube) or per rectum daily until discharge.

As a platform trial, and in a factorial design, patients could be simultaneously randomised to other treatment groups: i) azithromycin or colchicine or dimethyl fumarate versus usual care, ii) convalescent plasma or monoclonal antibody (REGN-CoV2) versus usual care, and iii) baricitinib versus usual care (appendix pp 30). Until 24 January 2021, the trial also allowed a subsequent randomisation for patients with progressive COVID-19 (evidence of hypoxia and a hyper-inflammatory state) to tocilizumab versus usual care. Participants and local study staff were not masked to the allocated treatment. The trial steering committee, investigators, and all other individuals involved in the trial were masked to aggregated outcome data during the trial.

### Procedures

A single online follow-up form was completed when participants were discharged, had died or at 28 days after randomisation, whichever occurred earliest (appendix p 35-41). Information was recorded on adherence to allocated study treatment, receipt of other COVID-19 treatments, duration of admission, receipt of respiratory or renal support, and vital status (including cause of death). In addition, in the UK, routine healthcare and registry data were obtained including information on vital status (with date and cause of death), discharge from hospital, receipt of respiratory support, or renal replacement therapy.

### Outcomes

Outcomes were assessed at 28 days after randomisation, with further analyses specified at 6 months. The primary outcome was all-cause mortality. Secondary outcomes were time to discharge from hospital, and, among patients not on invasive mechanical ventilation at randomisation, progression to invasive mechanical ventilation (including extra-corporeal membrane oxygenation) or death. Prespecified subsidiary clinical outcomes were use of non-invasive respiratory support, time to successful cessation of invasive mechanical ventilation (defined as cessation of invasive mechanical ventilation within, and survival to, 28 days), use of renal dialysis or haemofiltration, cause-specific mortality, major bleeding events (defined as intracranial bleeding or bleeding requiring transfusion, endoscopy, surgery or vasoactive drugs), thrombotic events (defined as acute pulmonary embolism, deep vein thrombosis, ischaemic stroke, myocardial infarction or systemic arterial embolism) and major cardiac arrhythmias. Information on suspected serious adverse reactions was collected in an expedited fashion to comply with regulatory requirements.

### Statistical Analysis

An intention-to-treat comparison was conducted between patients randomised to aspirin and patients randomised to usual care but for whom aspirin was both available and suitable as a treatment. For the primary outcome of 28-day mortality, the log-rank observed minus expected statistic and its variance were used to both test the null hypothesis of equal survival curves (i.e., the log-rank test) and to calculate the one-step estimate of the average mortality rate ratio. We constructed Kaplan-Meier survival curves to display cumulative mortality over the 28-day period. We used the same method to analyse time to hospital discharge and successful cessation of invasive mechanical ventilation, with patients who died in hospital right-censored on day 29. Median time to discharge was derived from Kaplan-Meier estimates. For the pre-specified composite secondary outcome of progression to invasive mechanical ventilation or death within 28 days (among those not receiving invasive mechanical ventilation at randomisation), and the subsidiary clinical outcomes of receipt of ventilation and use of haemodialysis or haemofiltration, the precise dates were not available and so the risk ratio was estimated instead.

Prespecified subgroup analyses (defined by characteristics at randomisation: age, sex, ethnicity, level of respiratory support, days since symptom onset, and use of corticosteroids) were performed for the primary outcome using the statistical test of interaction (test for heterogeneity or trend), in accordance with the prespecified analysis plan (appendix p 113). A sensitivity analysis restricting analysis of the primary outcome to patients with a positive PCR test for SARS-COV-2 was conducted. In addition, post-hoc exploratory analyses of the primary and secondary outcomes by venous thromboprophylaxis treatment at randomisation was conducted. Observed effects within subgroup categories were compared using a chi-squared test for heterogeneity or trend, in accordance with the prespecified analysis plan.

Estimates of rate and risk ratios are shown with 95% confidence intervals. All p-values are 2-sided and are shown without adjustment for multiple testing. The full database is held by the study team which collected the data from study sites and performed the analyses at the Nuffield Department of Population Health, University of Oxford (Oxford, UK).

As stated in the protocol, appropriate sample sizes could not be estimated when the trial was being planned at the start of the COVID-19 pandemic (appendix p 53). As the trial progressed, the trial steering committee, whose members were unaware of the results of the trial comparisons, determined that sufficient patients should be enrolled to provide at least 90% power at a two-sided significance level of 1% to detect a clinically relevant proportional reduction in 28-day mortality of 12.5% between the two groups. Consequently, on 21 March, 2021, the steering committee, masked to the results, closed recruitment to the aspirin comparison as sufficient patients had been recruited.

Analyses were performed using SAS version 9.4 and R version 4.0.3. The trial is registered with ISRCTN (50189673) and clinicaltrials.gov (NCT04381936).

## RESULTS

Between 1 November 2020 and 21 March 2021, 14892 (66%) of 22560 patients enrolled into the RECOVERY trial were eligible to be randomly allocated to aspirin (i.e. aspirin was available in the hospital at the time and the attending clinician was of the opinion that the patient had no known indication for or contraindication to aspirin, figure 1). 7351 patients were randomly allocated to usual care plus aspirin and 7541 were randomly allocated to usual care alone. The mean age of study participants in this comparison was 59.2 years (SD 14.2) and the median time since symptom onset was 9 days (IQR 6 to 12 days) (webtable 1). At randomisation, 5035 patients (34%) were receiving thromboprophylaxis with higher dose low molecular weight heparin (LMWH), 8878 (60%) with standard dose LMWH, and 979 (7%) were not receiving thromboprophylaxis.

**Figure 1:**
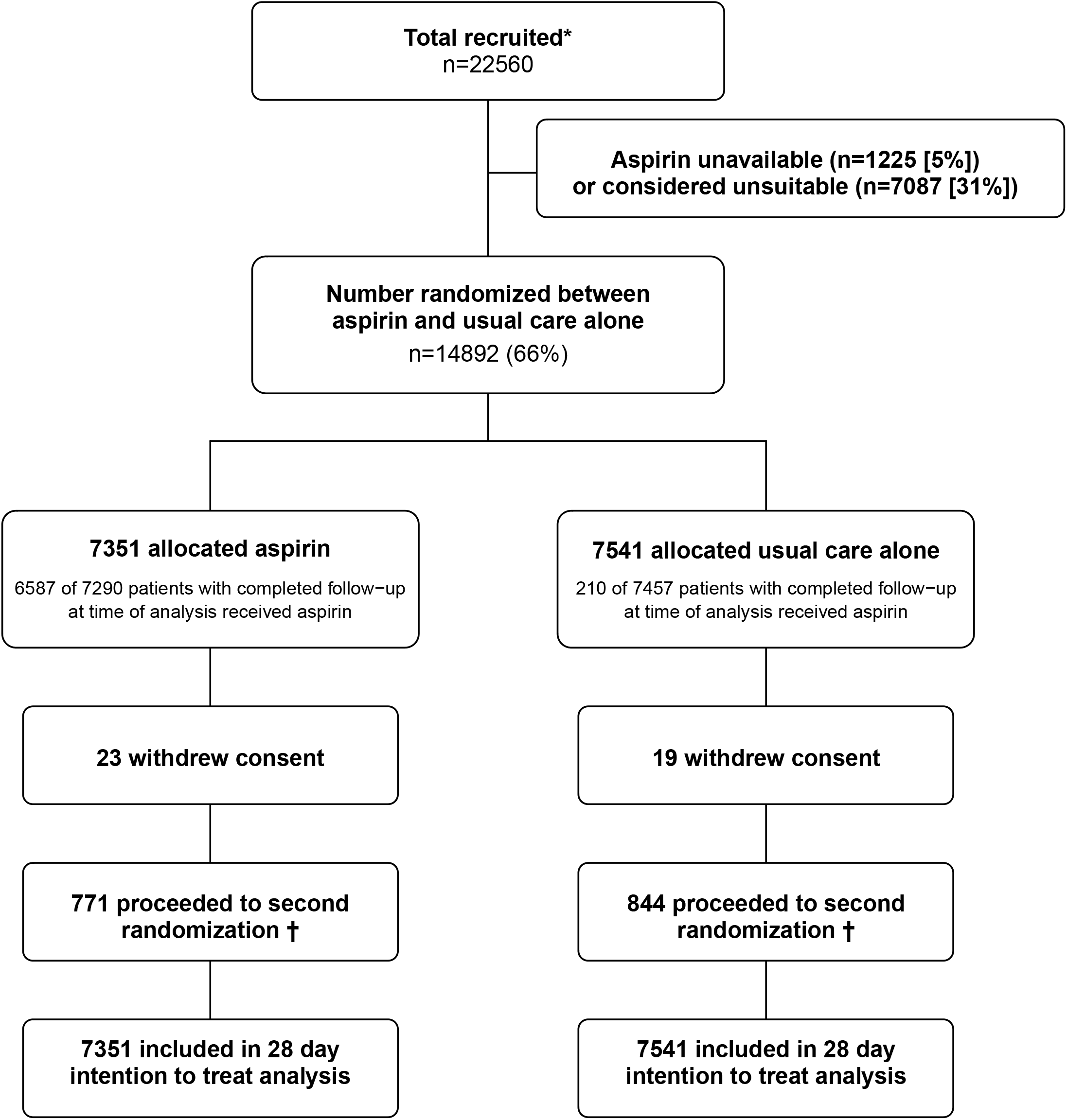
Trial profile. ITT=intention to treat. ^*^Number recruited overall during period that adult participants could be recruited into aspirin comparison. † Includes 379/7351 (5.2%) patients in the aspirin arm and 407/7541 (5.4%) patients in the usual care arm allocated to tocilizumab.

The follow-up form was completed for 7290 (99%) participants in the aspirin group and 7457 (99%) participants in the usual care group. Among participants with a completed follow-up form, 6587 (90%) allocated to aspirin received at least one dose and 210 (3%) allocated to usual care received at least one dose of aspirin (figure 1; webtable 2). Of the 6587 participants allocated to aspirin that received at least one dose of aspirin, 5040 (77%) received aspirin on most days following randomisation (≥90% of the days from randomisation to time to discharge or 28 days after randomisation, whichever was earlier). Use of other treatments for COVID-19 was similar among participants allocated aspirin and among those allocated usual care, with nearly 90% receiving a corticosteroid, about one-quarter receiving remdesivir, and one-eighth receiving tocilizumab (webtable 2).

Primary and secondary outcome data are known for 99% of randomly assigned patients. We observed no significant difference in the proportion of patients who met the primary outcome of 28-day mortality between the two randomised groups (1222 [17%] patients in the aspirin group vs. 1299 (17%) patients in the usual care group; rate ratio 0·96; 95% confidence interval [CI], 0·89 to 1·04; p=0·35; figure 2, table 2). The rate ratio was similar across all pre-specified sub-groups (figure 3). In an exploratory analysis restricted to the 14467 (97%) patients with a positive SARS-CoV-2 test result, the result was virtually identical (rate ratio 0.96, 95% CI 0·89 to 1·04; p=0·31).

**Figure 2:**
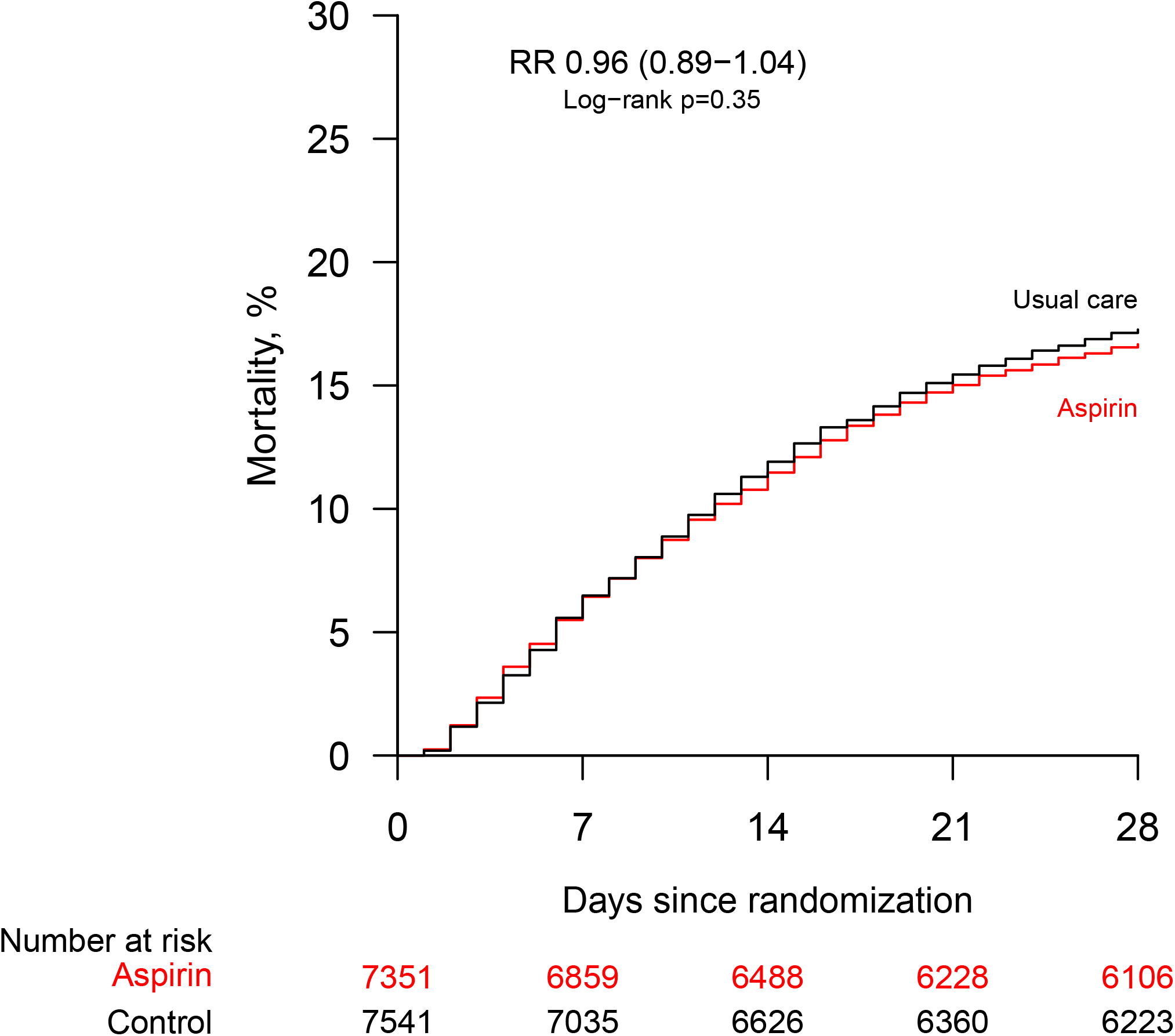
Effect of allocation to aspirin on 28-day mortality.

**Table 1:**
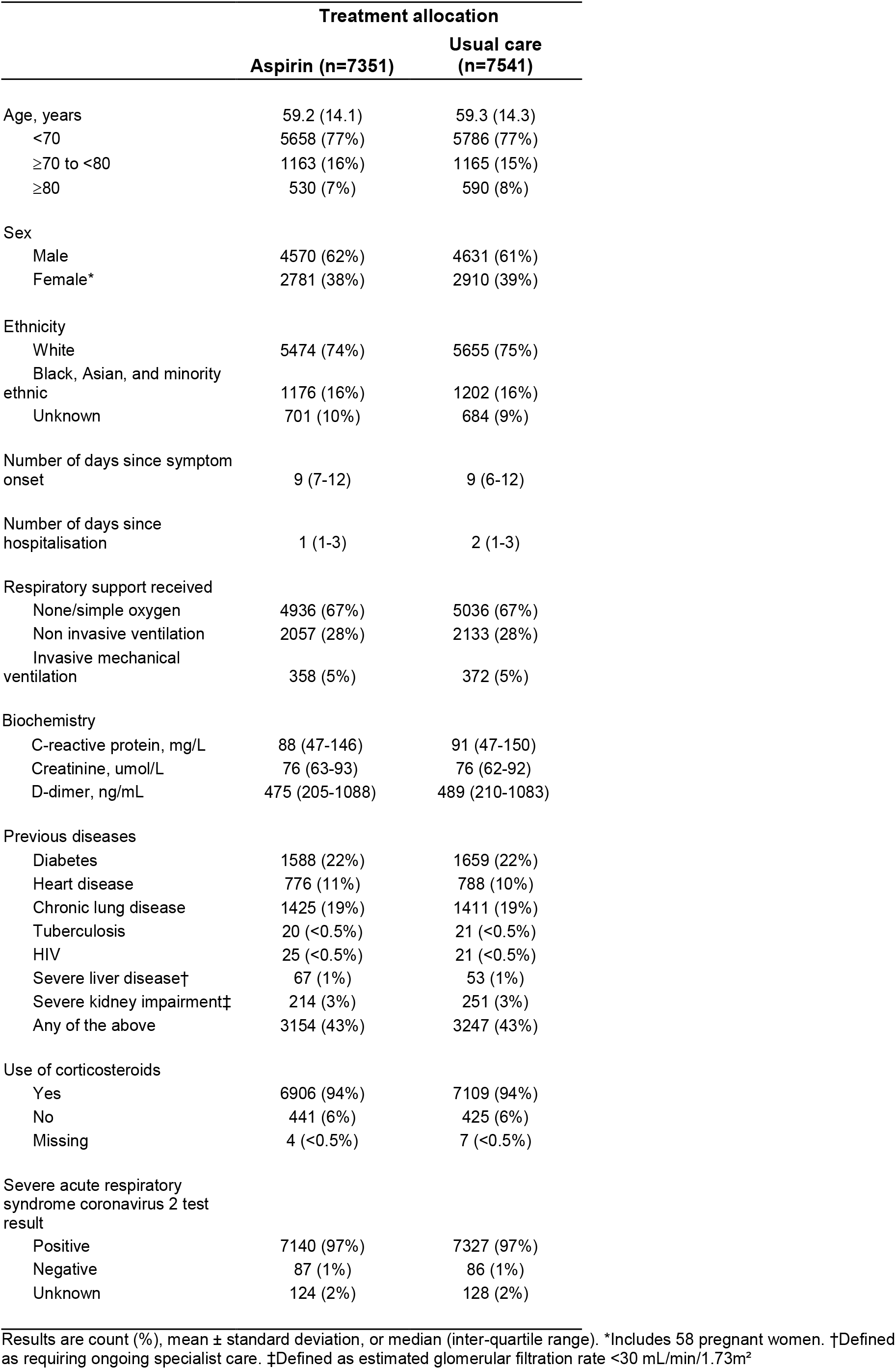
Baseline characteristics.

|  | Treatment allocation |  |
| --- | --- | --- |
|  | Aspirin (n=7351) | Usual care (n=7541) |
| Age, years | 59.2 (14.1) | 59.3 (14.3) |
| <70 | 5658 (77%) | 5786 (77%) |
| ≥70 to <80 | 1163 (16%) | 1165 (15%) |
| ≥80 | 530 (7%) | 590 (8%) |
| Sex |  |  |
| Male | 4570 (62%) | 4631 (61%) |
| Female* | 2781 (38%) | 2910 (39%) |
| Ethnicity |  |  |
| White | 5474 (74%) | 5655 (75%) |
| Black, Asian, and minority ethnic | 1176 (16%) | 1202 (16%) |
| Unknown | 701 (10%) | 684 (9%) |
| Number of days since symptom onset | 9 (7-12) | 9 (6-12) |
| Number of days since hospitalisation | 1 (1-3) | 2 (1-3) |
| Respiratory support received |  |  |
| None/simple oxygen | 4936 (67%) | 5036 (67%) |
| Non invasive ventilation | 2057 (28%) | 2133 (28%) |
| Invasive mechanical ventilation | 358 (5%) | 372 (5%) |
| Biochemistry |  |  |
| C-reactive protein, mg/L | 88 (47-146) | 91 (47-150) |
| Creatinine, umol/L | 76 (63-93) | 76 (62-92) |
| D-dimer, ng/mL | 475 (205-1088) | 489 (210-1083) |
| Previous diseases |  |  |
| Diabetes | 1588 (22%) | 1659 (22%) |
| Heart disease | 776 (11%) | 788 (10%) |
| Chronic lung disease | 1425 (19%) | 1411 (19%) |
| Tuberculosis | 20 (<0.5%) | 21 (<0.5%) |
| HIV | 25 (<0.5%) | 21 (<0.5%) |
| Severe liver disease† | 67 (1%) | 53 (1%) |
| Severe kidney impairment‡ | 214 (3%) | 251 (3%) |
| Any of the above | 3154 (43%) | 3247 (43%) |
| Use of corticosteroids |  |  |
| Yes | 6906 (94%) | 7109 (94%) |
| No | 441 (6%) | 425 (6%) |
| Missing | 4 (<0.5%) | 7 (<0.5%) |
| Severe acute respiratory syndrome coronavirus 2 test result |  |  |
| Positive | 7140 (97%) | 7327 (97%) |
| Negative | 87 (1%) | 86 (1%) |
| Unknown | 124 (2%) | 128 (2%) |

Results are count (%), mean ± standard deviation, or median (inter-quartile range). \*Includes 58 pregnant women. †Defined as requiring ongoing specialist care. ‡Defined as estimated glomerular filtration rate <30 mL/min/1.73m<sup>2</sup>

**Table 2:** Effect of allocation to aspirin on key study outcomes.

|  | Treatment allocation |  |  |  |
| --- | --- | --- | --- | --- |
|  | Aspirin (n=7351) | Usual care (n=7541) | RR (95% CI) | p value |
| Primary outcome: |  |  |  |  |
| 28-day mortality | 1222 (17%) | 1299 (17%) | 0.96 (0.89-1.04) | 0.35 |
| Secondary outcomes: |  |  |  |  |
| Median time to being discharged alive, days | 8 (5 to >28) | 9 (5 to >28) |  |  |
| Discharged from hospital within 28 days | 5496 (75%) | 5548 (74%) | 1.06 (1.02-1.10) | 0.0062 |
| Receipt of invasive mechanical ventilation or death* | 1473/6993 (21%) | 1569/7169 (22%) | 0.96 (0.90-1.03) | 0.23 |
| Invasive mechanical ventilation | 772/6993 (11%) | 829/7169 (12%) | 0.95 (0.87-1.05) | 0.32 |
| Death | 1076/6993 (15%) | 1141/7169 (16%) | 0.97 (0.90-1.04) | 0.39 |
| Subsidiary clinical outcomes |  |  |  |  |
| Use of ventilation | 1131/4936 (23%) | 1198/5036 (24%) | 0.96 (0.90-1.03) | 0.30 |
| Non-invasive ventilation | 1101/4936 (22%) | 1162/5036 (23%) | 0.97 (0.90-1.04) | 0.36 |
| Invasive mechanical ventilation | 296/4936 (6%) | 325/5036 (6%) | 0.93 (0.80-1.08) | 0.35 |
| Successful cessation of invasive mechanical ventilation | 135/358 (38%) | 135/372 (36%) | 1.08 (0.85-1.37) | 0.54 |
| Renal replacement therapy | 273/7291 (4%) | 282/7480 (4%) | 0.99 (0.84-1.17) | 0.93 |
| RR=Rate Ratio for the outcomes of 28-day mortality and hospital discharge, and risk ratio for the outcome of receipt of invasive mechanical ventilation or death (and its subcomponents). CI=confidence interval. *Analyses exclude those on invasive mechanical ventilation at randomization. |  |  |  |  |

**Figure 3:**
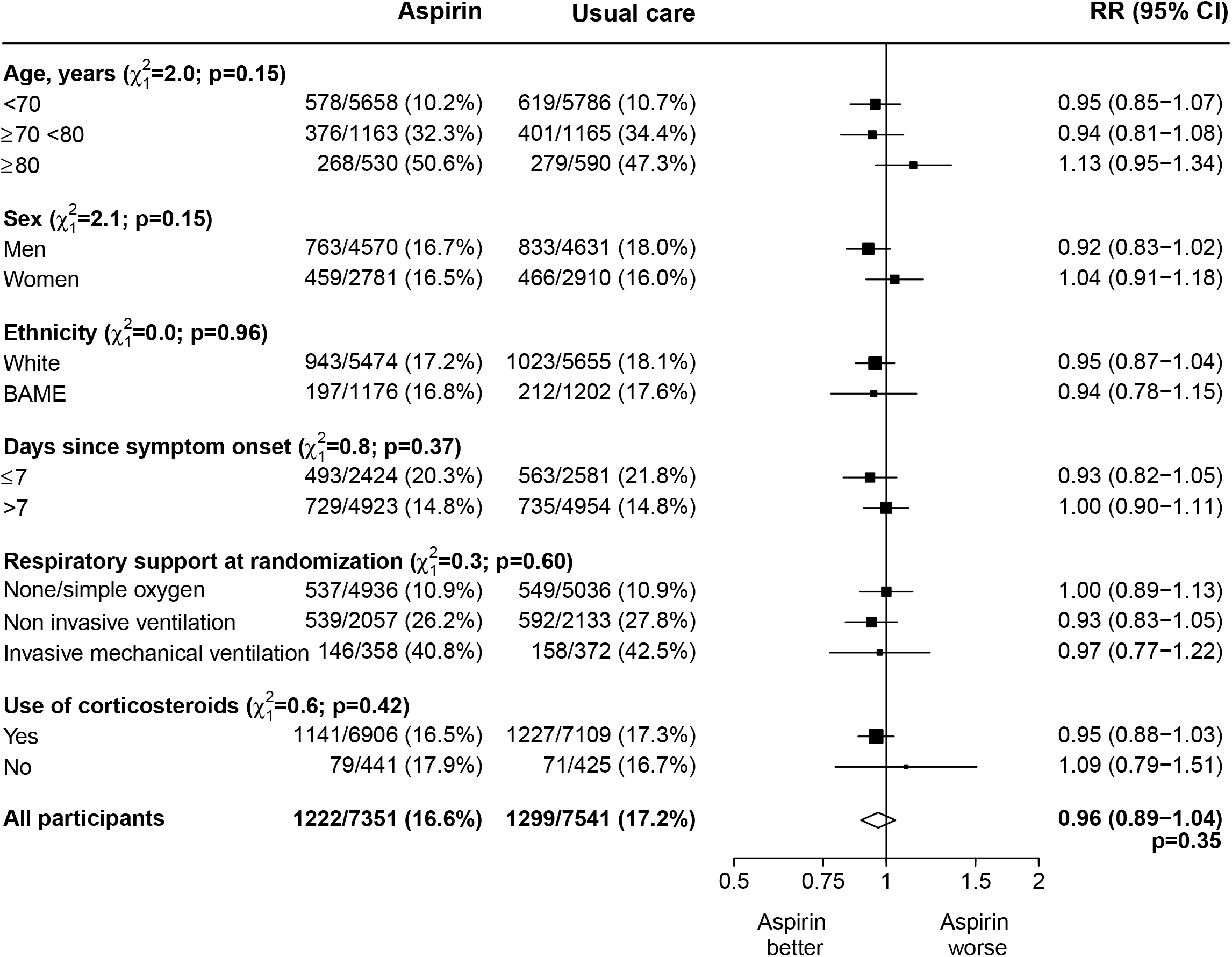
Effects of allocation to aspirin on 28-day mortality by baseline characteristics. Subgroup–specific rate ratio estimates are represented by squares (with areas of the squares proportional to the amount of statistical information) and the lines through them correspond to the 95% CIs. The ethnicity, days since onset and use of corticosteroids subgroups exclude those with missing data, but these patients are included in the overall summary diamond.

Allocation to aspirin was associated with a reduction of 1 day in median time until discharge alive from hospital compared to usual care (median 8 days vs. 9 days [IQR for each 5 to >28 days]) and an increased rate of discharge alive within 28 days (75% vs. 74%, rate ratio 1·06, 95% CI 1·02 to 1·10, p=0·0062) (table 2). Among those not on invasive mechanical ventilation at baseline, the number of patients progressing to the pre-specified composite secondary outcome of invasive mechanical ventilation or death among those allocated to aspirin was similar to that among those allocated to usual care (21% vs. 22%, risk ratio 0·96, 95% CI 0·90 to 1·03, p=0·23). There was no evidence that the effect of allocation to aspirin vs. usual care on time until discharge alive from hospital or on invasive mechanical ventilation or death differed between the pre-specified subgroups of patients (webfigure 1, webfigure 2). In a post-hoc exploratory analysis there was no evidence that the effect of allocation to aspirin vs. usual care on the primary and secondary outcomes differed by use of LMWH use at randomisation (webfigure 3).

We found no significant differences in the prespecified subsidiary clinical outcomes of cause-specific mortality (webtable 3), use of ventilation, successful cessation of invasive mechanical ventilation, or receipt of renal dialysis or haemofiltration (table 2). As expected with the use of aspirin, the incidence of thrombotic events was lower (4.6% vs. 5.3%; absolute difference 0.6%, SE 0.4%) and the incidence of major bleeding events was higher (1.6% vs. 1.0%; absolute difference 0.6%, SE 0.2%) in the aspirin group (webtable 4). The incidence of new cardiac arrhythmias was similar in the two groups (webtable 5). There were 18 reports of a serious adverse event believed related to aspirin, all of which were due to haemorrhagic events (webtable 6).

## DISCUSSION

In this large, randomised trial involving over 14,000 patients and over 2000 deaths, allocation to aspirin was not associated with reductions in mortality or, among those not on invasive mechanical ventilation at baseline, the risk of progressing to the composite endpoint of invasive mechanical ventilation or death. Allocation to aspirin was, however, associated with a small increase in the rate of being discharged from hospital alive within 28 days. These results were consistent across the prespecified subgroups of age, sex, ethnicity, duration of symptoms prior to randomisation, level of respiratory support at randomisation, and use of corticosteroids.

As expected, allocation to aspirin was associated with an increased risk of major bleeding and a decreased risk of thromboembolic complications, such that for every 1000 patients treated with aspirin, approximately 6 more would experience a major bleeding event and approximately 6 fewer would experience a thromboembolic event. The rate of reported thromboembolic events in our study population was low (5.3% in the usual care arm) in comparison with previous reports.^1,2^ This could be related to the widespread use of corticosteroids in the trial population resulting in reduced thrombo-inflammatory stimulus or because of the exclusion of patients already receiving aspirin because of prior cardiovascular disease. It is possible that aspirin might have a more meaningful benefit in populations with a higher thrombotic risk, although there would also likely be a corresponding increase in bleeding risk.^15^

The pathogenesis of thromboembolism in COVID-19 is likely to be multifactorial. Coagulopathy is common in severe COVID-19 and is associated with an inflammatory state, neutrophil extracellular traps, and poor outcomes.^2,16-20^ Platelet activation is increased as a result (and potentially by direct interaction with the virus), amplifying inflammation locally and triggering immunothrombosis.^21,22^ In addition, SARS-CoV-2 infection can cause inflammation, dysfunction, and disruption of the vascular endothelium in multiple organs, potentially via direct entry through the ACE-2 receptor.^23-25^ The resulting endothelial injury and tissue factor exposure promote thrombosis in the pulmonary circulation and other vascular beds, with microangiopathy and alveolar capillary occlusion contributing to the diffuse alveolar damage and hypoxemia seen in COVID-19.^24,26^ Furthermore, in autopsy studies pulmonary microthrombi are nine times more frequent in patients with COVID-19 compared to patients with influenza.^24^

A large number of randomised controlled trials of antithrombotic therapy in COVID-19 are registered, including trials of therapeutic doses of heparin, direct acting oral anticoagulants, anti-platelet agents, serine protease inhibitors, and thrombolytics.^27^ In critically-ill patients, the INSPIRATION, REMAP-CAP, ACTIV-4a and ATTACC trials did not report a benefit in clinical outcomes from therapeutic anticoagulation.^28,29^ Similarly, preliminary results from the COALIZAO-ACTION trial did not show a benefit from therapeutic anticoagulation (either heparin or rivaroxaban) in a combined endpoint of mortality, successful discharge, or need for oxygen in hospitalised patients with elevated D-dimers.^30^ However, the REMAP-CAP/ACTIV-4a/ATTACC investigators have reported that in non-critically ill COVID-19 patients, compared to thromboprophylaxis doses, heparin at therapeutic doses was associated with an absolute increase of 4.6% (95% credible interval 0.7 to 8.1) in the proportion of participants surviving to hospital discharge without receipt of organ support during the first 21 days.^31^

Although there are currently no other published randomised trial data on the use of aspirin in COVID-19, the REMAP-CAP/ACTIV-4a/ATTACC report does suggest that antithrombotic therapy may be important in some patients.^31^ The lack of meaningful benefit from aspirin in our trial could be because antiplatelet therapy confers no significant additional benefit on top of high rates of antithrombotic therapy with LMWH and corticosteroid treatment diminishing thrombo-inflammatory stimulation. Alternatively, other non-platelet pathways leading to thrombosis and alveolar damage may be more important determinants of clinical outcomes.

Any potential benefit of antithrombotic therapies in COVID-19 patients may also depend on timing of treatment initiation, especially if thrombi have already developed by the time of admission.^32^ Thromboembolic events and microthrombi are common in COVID-19 patients on either prophylactic or therapeutic anticoagulation.^33^ The apparent lack of benefit in INSPIRATION and the REMAP-CAP/ACTIV-4a/ATTACC severe disease cohorts suggests that these patients might have passed the point at which any benefit from therapeutic anticoagulation could be gained.^28,29^ Although we found no evidence of heterogeneity based on duration of symptoms, baseline disease severity, or background thrombotic prophylaxis regimen, ongoing trials of aspirin in ambulatory populations and those exploring more potent anti-platelet inhibition and fibrinolysis should provide further insights.

Strengths of this trial included that it was randomised, had a large sample size, broad eligibility criteria, and 99% of patients were followed up for the primary outcome. The trial also had some limitations. Detailed information on radiological or physiological outcomes was not collected. Although this randomised trial is open label (i.e., participants and local hospital staff are aware of the assigned treatment), the primary and secondary outcomes are unambiguous and were ascertained without bias through linkage to routine health records. However, it cannot be excluded that reporting of thromboembolic and bleeding events might have been influenced by knowledge of treatment allocation. Nevertheless, the proportional effects of aspirin on these events were very similar to those reported in previous large clinical trials of aspirin in people with prior cardiovascular disease.^6^

The RECOVERY trial only studied hospitalised COVID-19 patients and, therefore, is not able to provide evidence on the safety and efficacy of aspirin used in other patient groups. Further studies to identify the safety and efficacy of aspirin in non-hospitalised patients are needed and are ongoing.

In summary, the results of this large, randomised trial do not support the addition of aspirin to standard thromboprophylaxis or therapeutic anticoagulation in patients hospitalised with COVID-19.

## Supporting information

CONSORT Checklist

## Data Availability

The protocol, consent form, statistical analysis plan, definition & derivation of clinical characteristics & outcomes, training materials, regulatory documents, and other relevant study materials are available online at www.recoverytrial.net. As described in the protocol, the trial Steering Committee will facilitate the use of the study data and approval will not be unreasonably withheld. Deidentified participant data will be made available to bona fide researchers registered with an appropriate institution within 3 months of publication. However, the Steering Committee will need to be satisfied that any proposed publication is of high quality, honours the commitments made to the study participants in the consent documentation and ethical approvals, and is compliant with relevant legal and regulatory requirements (e.g. relating to data protection and privacy). The Steering Committee will have the right to review and comment on any draft manuscripts prior to publication. https://www.ndph.ox.ac.uk/data-access

## Contributors

This manuscript was initially drafted by PWH and MJL, further developed by the Writing Committee, and approved by all members of the trial steering committee. PWH and MJL vouch for the data and analyses, and for the fidelity of this report to the study protocol and data analysis plan. PWH, JKB, MB, LCC, JD, SNF, TJ, EJ, KJ, WSL, AM, AM, KR, GT, MM, RH, and MJL designed the trial and study protocol. MM, MC, G P-A, LP, NB, ST, VC, TB, HT, BE, DC, TW, RS, CG and the Data Linkage team at the RECOVERY Coordinating Centre, and the Health Records and Local Clinical Centre staff listed in the appendix collected the data. NS, ES and JRE did the statistical analysis. All authors contributed to data interpretation and critical review and revision of the manuscript. PWH and MJL had access to the study data and had final responsibility for the decision to submit for publication.

## Data Monitoring Committee

Peter Sandercock, Janet Darbyshire, David DeMets, Robert Fowler, David Lalloo, Mohammed Munavvar (from January 2021), Ian Roberts (until December 2020), Janet Wittes.

## Data sharing

The protocol, consent form, statistical analysis plan, definition & derivation of clinical characteristics & outcomes, training materials, regulatory documents, and other relevant study materials are available online at www.recoverytrial.net. As described in the protocol, the trial Steering Committee will facilitate the use of the study data and approval will not be unreasonably withheld. Deidentified participant data will be made available to bona fide researchers registered with an appropriate institution within 3 months of publication. However, the Steering Committee will need to be satisfied that any proposed publication is of high quality, honours the commitments made to the study participants in the consent documentation and ethical approvals, and is compliant with relevant legal and regulatory requirements (e.g. relating to data protection and privacy). The Steering Committee will have the right to review and comment on any draft manuscripts prior to publication. Data will be made available in line with the policy and procedures described at: https://www.ndph.ox.ac.uk/data-access. Those wishing to request access should complete the form at https://www.ndph.ox.ac.uk/files/about/data_access_enquiry_form_13_6_2019.docx and e-mail to

## Acknowledgements

Above all, we would like to thank the thousands of patients who participated in this trial. We would also like to thank the many doctors, nurses, pharmacists, other allied health professionals, and research administrators at 176 NHS hospital organisations across the whole of the UK, supported by staff at the National Institute of Health Research (NIHR) Clinical Research Network, NHS DigiTrials, Public Health England, Department of Health & Social Care, the Intensive Care National Audit & Research Centre, Public Health Scotland, National Records Service of Scotland, the Secure Anonymised Information Linkage (SAIL) at University of Swansea, and the NHS in England, Scotland, Wales and Northern Ireland.

The RECOVERY trial is supported by grants to the University of Oxford from UK Research and Innovation (UKRI)/NIHR (Grant reference: MC_PC_19056), the Department of Health and Social Care (DHSC)/UKRI/NIHR COVID-19 Rapid Response Grant (COV19-RECPLA) and the Wellcome Trust (Grant Ref: 222406/Z/20/Z) through the COVID-19 Therapeutics Accelerator. Core funding is provided by NIHR Oxford Biomedical Research Centre, Wellcome, the Bill and Melinda Gates Foundation, the Foreign, Commonwealth and development Department, Health Data Research UK, the Medical Research Council Population Health Research Unit, the NIHR Health Protection Unit in Emerging and Zoonotic Infections, and NIHR Clinical Trials Unit Support Funding. TJ is supported by a grant from UK Medical Research Council (MC_UU_00002/14) and an NIHR Senior Research Fellowship (NIHR-SRF-2015-08-001). WSL is supported by core funding provided by NIHR Nottingham Biomedical Research Centre. Combiphar supplied colchicine free of charge for use in this trial in Indonesia. Abbvie contributed some supplies of lopinavir-ritonavir for use in this trial. Tocilizumab was provided free of charge for this trial by Roche Products Limited. REGN-COV2 was provided free of charge for this trial by Regeneron.

The views expressed in this publication are those of the authors and not necessarily those of the NHS or the NIHR.

